# Submicroscopic burden of zoonotic *P. knowlesi* malaria on Mursala Island and *P. falciparum* and *P. vivax* transmission in mainland North Sumatra, Indonesia

**DOI:** 10.1101/2025.06.02.25328766

**Authors:** Inke ND Lubis, Ranti Permatasari, Lambok Siahaan, R Andika Dwi Cahyadi, Irbah Rhea Alvieda Nainggolan, Rycha Dwi Syafutri, Monica Nadya Sinambela, Silvia Jauharah, Agatha Lestari, Minerva Theodora, Hellen Prameswari, Kim A Piera, Bridget E Barber, Nicholas M Anstey, Matthew J Grigg

## Abstract

Accurate molecular tools are essential for estimating zoonotic malaria transmission in Southeast Asia. This study applied ultrasensitive reverse-transcriptase real-time PCR to detect zoonotic malaria in febrile patients from eight mainland health facilities and Mursala Island, spanning three districts in North Sumatra, Indonesia. Among 64 participants on Mursala, 7 (10.9%) had confirmed *Plasmodium knowlesi* symptomatic infections, including two submicroscopic infections associated with severe anaemia. All were negative by microscopy and pan-pLDH rapid diagnostic tests. No *P. knowlesi* infections were identified among 947 participants from mainland sites; PCR detected malaria in 34%, including *P. vivax* (17.5%) and *P. falciparum* (7.5%). Of these, 30% were submicroscopic infections. No *P. cynomolgi* infections were identified. *P. knowlesi* transmission is low in North Sumatra, however, may cause serious disease. Molecular diagnostics remain crucial for identifying zoonotic malaria and should be integrated into surveillance systems to inform public health control measures.

## Introduction

Zoonotic malaria caused by *Plasmodium knowlesi* has emerged as a significant public health challenge in Southeast Asia^1,2^. Indonesia has a sizeable population-at-risk of zoonotic malaria^3,4^ due to its ecological diversity and widespread overlap between the parasite’s major natural macaque hosts (*Macaca fascicularis* and *M. nemestrina*) and *Anopheles leucosphyrus* group vectors^5^ . Rapid agricultural expansion and land use changes have amplified the risk of zoonotic malaria by increasing interaction between humans, macaques, and mosquito vectors^6,7^. However, understanding of *P. knowlesi* transmission in Indonesia is limited due to being frequently misdiagnosed as non-zoonotic species using routine microscopy, with *P. falciparum* and *P. vivax* often incorrectly identified even more so than the morphologically similar *P. malariae*^8,9^. Lateral flow-based rapid diagnostic tests (RDTs), designed and commonly used for detection of *P. falciparum* and *P. vivax*, have also lacked the specificity, and until recently the sensitivity, required for reliable *P. knowlesi* identification, particularly at low parasite densities^10–12^. Diagnostic misclassification using available point-of-care diagnostics has previously obscured the actual burden of *P. knowlesi* malaria, a trend first observed in Sarawak, Malaysian Borneo, where a rise in cases of microscopy-diagnosed *P. malariae* ultimately led to the initial molecular identification of *P. knowlesi* as a dominant cause of human malaria in 2004^13^ .

The use of accurate PCR assays in research settings have highlighted the presence of *P. knowlesi* transmission across western Indonesia, particularly within the provinces of Sumatra^8,9,14–19^ and Kalimantan^20–24^, but outside the non-endemic eastern province of Papua where the highest burden of non-zoonotic malaria remains^25^. However, there remain significant gaps in our understanding of the distribution of *P. knowlesi* within areas such as North Sumatra, where diverse ecological conditions are conducive to zoonotic malaria transmission^3^ but where many districts have not been systematically evaluated using sensitive molecular tools. *P. knowlesi* transmission has been reported to date in the districts of Batubara, Langkat, Dairi and the islands of South Nias^9,18,26^. Other zoonotic malaria species such as *P. cynomolgi* have not been screened for in western Indonesia despite sharing the same macaque hosts and mosquito vectors^27^. Understanding the prevalence and distribution of zoonotic *Plasmodium* species in regions like North Sumatra is critical for improving malaria control efforts, meeting WHO malaria elimination goals, and addressing the hidden burden of zoonotic malaria in Indonesia^28^ . The aim of this study was to conduct molecular epidemiological surveillance using improved ultrasensitive PCR for both zoonotic *P. knowlesi* and *P. cynomolgi* detection in high-risk areas across North Sumatra.

## Methods

### Study design

The main study design consisted of longitudinal passive malaria case detection of febrile patient presentations at selected health facilities on mainland North Sumatra, Indonesia. A separate small cross-sectional study was conducted on Mursala Island.

### Study sites

The health facility surveys on the North Sumatra mainland covered a study area of 6,933 km^2^ with an estimated 1,579,000 people at-risk^29^ (**Figure 1**). There were 8 health facilities from across 3 districts, including: 4 government primary health clinics in Batubara, 2 primary health clinics and a referral hospital in Central Tapanuli, and an urban referral hospital in Tanjung Balai. The estimated prevalence of malaria was taken into account when selecting the health facility locations; at the time of study design in 2019 the annual malaria provincial-level malaria cases reported included 41% from Batubara, 10% in Asahan and 2% in Central Tapanuli^1,3^.

**Figure 1.**
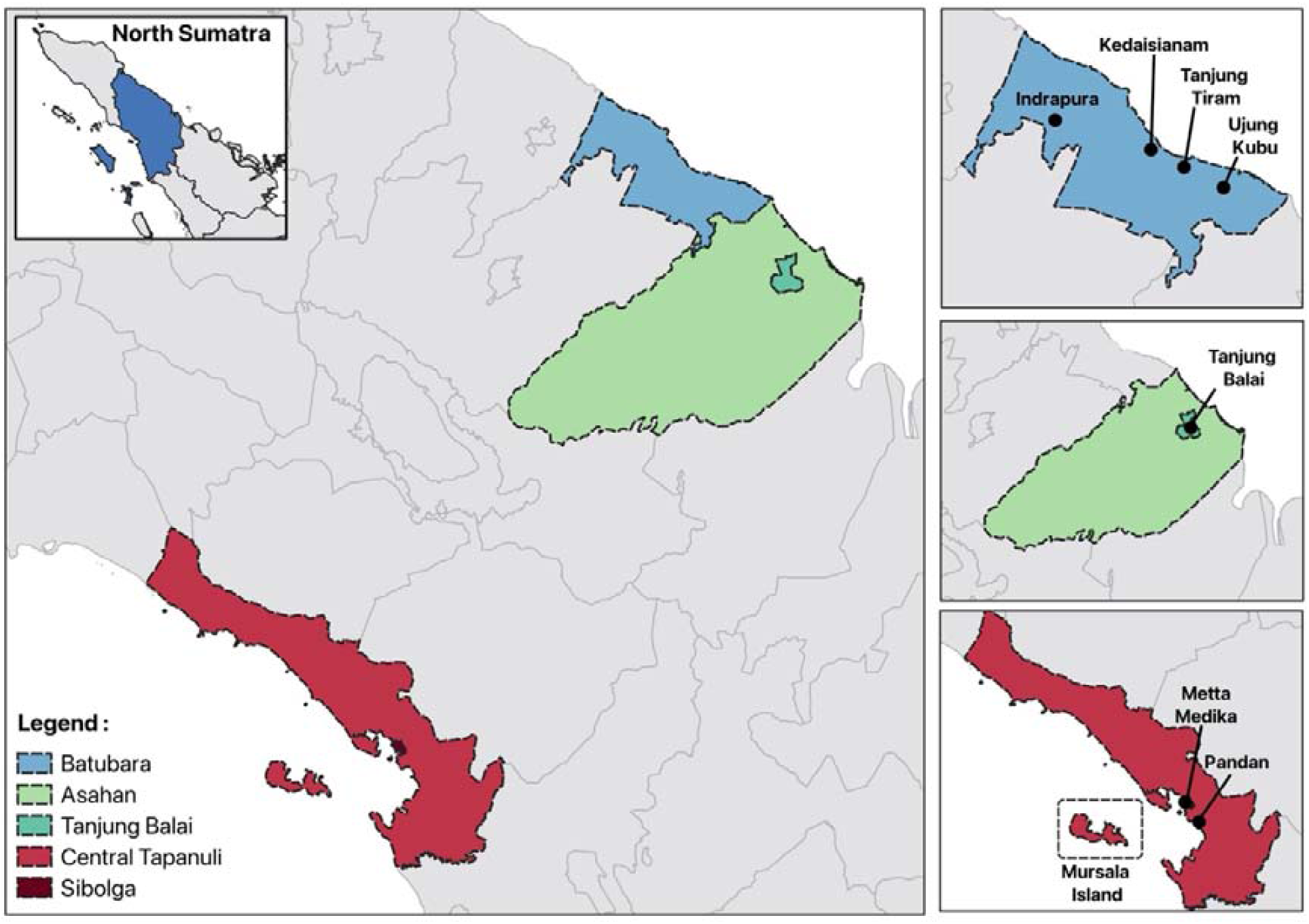
Map of study sites: Batubara district (blue), Tanjung Balai district (green); and Central Tapanuli (red).

A separate cross-sectional study was performed on Mursala Island, also part of Central Tapanuli district located 22km off the coast and inhabited by 5,400 people with a size of 80 km^2^. Community approval and awareness for the survey was obtained after meeting with village leaders and heads of households.

### Study participants

Patients at mainland study health facilities were enrolled if they met the following inclusion criteria: age of at least 1 year, body temperature above 37.5LC or a history of fever in last 48 hours, presented with non-specific symptoms (suspected malaria), resided primarily in the study district within previous 1 year, and provided informed written consent (from parents/guardians if less than 18 years old). For Mursala Island, criteria were similar although a history of fever was not required. Demographic, clinical, and epidemiological data were collected by trained research staff using a standardised electronic case record form (REDCap, version 10.6). Severe malaria was defined using WHO 2014 Research and Epidemiological criteria for severe disease^30^ .

### Blood sampling

Venous whole blood (or capillary sample in those ≤10 years old and all participants from Mursala) was collected in ethylenediaminetetraacetic acid (EDTA) for standard haematology and biochemistry at local laboratories where available. Haemoglobin was measured with a HemoCue® 201 machine (HemoCue AB, Sweden) on Mursala Island. Malaria screening was performed using Ministry of Health supplied routine malaria rapid diagnostic test (RDTs) with combined PAN-*Plasmodium* species parasite lactate dehydrogenase (PAN-pLDH) and *P. falciparum*-specific histidine-rich-protein-2 (Pf-HRP2) targets depending on site availability, and microscopic examination of blood smears stained with Giemsa. Slides and RDTs were read by local health facility officers, and slides were re-examined and quantified by an experienced microscopist blinded to the first microscopy and RDT results according to standard WHO procedures^31^. A total of 300μl and 100μl of whole blood from adults and children, respectively, were placed in field-stable DNA/RNA Shield^TM^ (Zymo Research, USA) total nucleic acid preservation media and frozen at -20°C prior to shipping to the Faculty of Medicine, Universitas Sumatera. Patients confirmed as malaria positive on point-of-care testing were treated according to national guidelines^32^.

### Laboratory malaria PCR detection

Total nucleic acids were extracted (QIAamp DNA Blood Mini Kit) from 200μL of the blood sample preserved in DNA/RNA Shield, followed by high-capacity cDNA reverse transcription (Applied Biosystems, USA). Amplification of cDNA was conducted using a quantitative real-time PCR (qRT-PCR) workflow targeting 18S rRNA genes^33^. Positive *Plasmodium* genus results were defined as Ct values below 40 from duplicate runs with a Ct value difference of less than 3. Those positive had conventional species-specific PCR assays performed using the same cDNA for *P. knowlesi*^34^ , *P. cynomolgi*^35^ and other human *Plasmodium* species (*P. falciparum, P. vivax, P. ovale spp.*, and *P. malariae*^36^) detection. Each PCR amplification included a *Plasmodium* species positive and negative control and molecular weight standards.

### Statistical analysis

All statistical analyses were performed using Stata version 17.0 (StataCorp, Texas, USA). Chi-squared or Fisher’s exact were used to evaluate proportional differences in binary variables, and Student’s t-test or Wilcoxin ranksum for pairwise comparisons of clinical and epidemiological data. Results of microscopy and RDT assays were evaluated against reference PCR, enabling calculation of diagnostic sensitivity and specificity with exact binomial 95% confidence intervals. The crude malaria incidence rate was calculated as the number of malaria cases per 100,000 at-risk people per year at a district catchment level. Study site district catchment populations were projected estimates from 2020 census data^37^. Logistic regression models were used for univariable and multivariable epidemiological associations with individual *Plasmodium* species infections, with odds ratios and 95% confidence intervals reported.

### Ethical approval

The study ethics approval was received from the medical research ethics committee of Universitas Sumatera Utara (No. 179/TGL/KEPK FK-USU-RSUPHAM/2019).

## Results

### Mainland health facility surveys

From August 2019 to December 2020, there were 3,377 patients with acute febrile illness presenting to the health facilities in mainland North Sumatra (**Figure 2**). Of these, 947 (28.1%) were enrolled including 704 (74.3%) patients from Batubara, 136 (14.4%) patients from Tanjung Balai, and 107 (11.3%) patients from Central Tapanuli. The median age was 17 years (IQR 12-34; range 2-67) and 48% were female. Overall, 321 (33.9%; 95%CI 30.9-37.0%) patients tested positive for malaria by PCR: 71 (7.5%) *P. falciparum*, 166 (17.5%) *P. vivax*, 5 (0.5%) mixed *P. falciparum*/*P. vivax*, and 79 (8.3%) where the exact *Plasmodium* species infection was unable to be determined. There were 96 febrile patients with submicroscopic infections (29.9% of total infections; 95% CI 24.9-35.2%), including 8 *P. falciparum*, 16 *P. vivax* and 72 *Plasmodium* genus.

**Figure 2:**
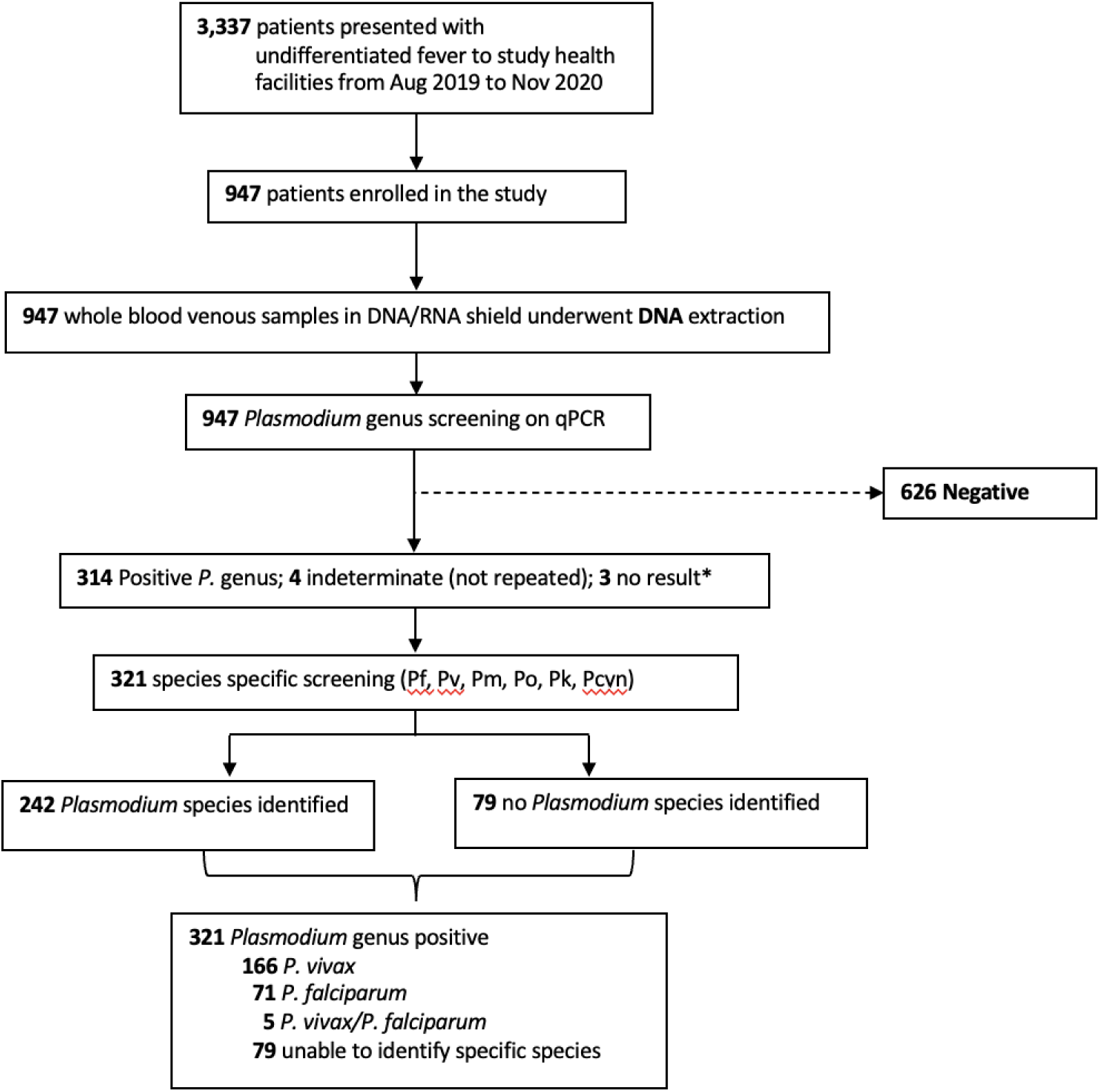
Enrolment flowchart for health facility surveys and *Plasmodium* species results.

There were no significant differences in the median age (∼16 years), or the proportion of children (∼35%) with *P. falciparum* or *P. vivax* infections (**Table 1**). The majority presented with non-specific symptoms, most commonly headache, nausea/vomiting, cough, and abdominal pain, with a median preceding fever duration of 6 days. Anaemia (using WHO age and sex haemoglobin criteria^38^) was observed in over 40% of patients infected with *P. falciparum*, *P. vivax* or undetermined *Plasmodium* species. Thrombocytopenia (platelet count <150×10^3^/μL) was present in over 75% of patients with *P. falciparum* or *P. vivax* malaria. In contrast, anaemia and thrombocytopenia were only present in 30% and 22% of the febrile non-malaria controls, respectively (*p*≤0.008). A previous history of malaria was positively associated with acute infections (OR 2.91, 95%CI 1.88-4.50, *p*<0.001).

**Table 1.** Clinical and laboratory features of *Plasmodium* species infections in mainland North Sumatra.

| Patient characteristic | North Sumatra – mainland sites |  |  |  |  |  |
| --- | --- | --- | --- | --- | --- | --- |
|  | *P. falciparum* | *P. vivax* | *P. falciparum* /*P. vivax* | *Plasmodium* genus | Control (negative) | *P*-value (across groups) |
| Number enrolled | 71 | 166 | 5 | 79 | 626 |  |
| Age, median years | 16 | 16 | 18 | 15 | 18 | 0.063 |
| IQR | 12 | 13 | 14 | 17 | 26 |  |
| Range | 2 - 55 | 4 - 65 | 4 - 30 | 2 - 54 | 1 - 80 |  |
| Children (age <15 yrs), n (%) | 25 (35.2) | 58 (34.9) | 2 (40) | 37 (46.8) | 229 (36.6) | 0.46 |
| Male gender, n (%) | 38 (53.5) | 100 (60.2) | 4 (80) | 33 (41.8) | 321 (51.3) | **0.035** |
| Previous malaria (self-reported), n (%) | 15 (21.1) | 31 (18.7) | 2 (40) | 5 (6.3) | 40 (6.4) | **<0.001** |
| Symptoms on enrolment, n (%) |  |  |  |  |  |  |
| Headache | 70 (98.6) | 147 (88.6) | 4 (80) | 62 (78.5) | 503 (80.4) | **<0.001** |
| Nausea | 54 (76.1) | 127 (76.5) | 4 (80) | 51 (64.6) | 422 (67.4) | 0.069 |
| Cough | 41 (57.7) | 75 (45.2) | 2 (40) | 40 (50.6) | 324 (51.8) | 0.45 |
| Shortness of breath | 11 (15.5) | 17 (10.2) | 0 (0) | 15 (18.9) | 140 (22.4) | **0.005** |
| Vomiting | 37 (52.1) | 75 (45.2) | 2 (40) | 29 (36.7) | 276 (44.1) | 0.432 |
| Abdominal pain | 36 (50.7) | 60 (36.1) | 4 (80) | 39 (49.4) | 313 (50) | **0.014** |
| Diarrhoea | 8 (11.3) | 17 (10.2) | 0 (0) | 6 (7.6) | 68 (10.9) | 0.921 |
| Bleeding | 0 (0) | 5 (3.0) | 0 (0) | 1 (1.3) | 4 (0.6) | 0.106 |
| Convulsions | 10 (14.1) | 13 (7.8) | 0 (0) | 4 (5.1) | 118 (18.8) | **<0.001** |
| Fever (temp >37.5°C), n (%) | 35 (49.3) | 101 (60.8) | 2 (40) | 51 (64.6) | 293 (46.8) | **0.001** |
| Febrile days (prior to presentation), median (IQR, range) | 6 (4, 1-40) | 6 (4, 1-30) | 7 (7, 5-15) | 5 (3, 2-25) | 5 (4, 0-40) |  |
| Microscopy result <sup>†</sup> |  |  |  |  |  |  |
| *P. vivax* | - | 103 | 2 | - | 11 | - |
| *P. falciparum* | 42 | 2 | 1 | 2 | 10 | - |
| *P. falciparum* / *P. vivax* | 20 | 44 | 2 | 3 | 6 | - |
| *P. falciparum* / *P. knowlesi* | 1 | - | - | - | - | - |
| *P. knowlesi* / *P. malariae* | - | 1 | - | - | - | - |
| Negative | 8 | 16 | - | 72 | 599 | - |
| Sensitivity, % (95%CI) | 59.2 (46.8-70.7) | 62.0 (54.2-69.5) | 40.0 (5.3-85.3) | - | 95.7 (93.8-97.1) |  |
| Specificity, % (95%CI) | 98.4 (97.3-99.1) | 92.4 (90.3-94.1) | 92.3 (90.4-93.9) |  | 70.1 (64.8-75.1) |  |
| Parasite count |  |  |  |  |  |  |
| Positive result, n (%) | 63 (89) | 152 (92) | 5 (100) | 8 (10) | - |  |
| Parasites/μL, geom. mean (95% CI) | 2844 (2055-3936) | 2273 (2487-3187) | 4597 (2781-7599) | 2211 (743-6577) | - |  |
| Parasites/μL, median | 3037 | 2200 | 5520 | 1780 | - |  |
| IQR | 1680-5502 | 1420-3200 | 3280-6280 | 880-5480 | - |  |
| Range | 28-31760 | 512-14560 | 2720-6640 | 480-23840 | - |  |
| RDT result<sup>†</sup> |  |  |  |  |  |  |
| (PAN-pLDH / Pf-HRP2) |  |  |  |  |  |  |
| Number conducted, n (%) | 51 (72) | 123 (74) | 3 (60) | 72 (91) | 441 (71) |  |
| Sensitivity, % (95%CI) | 82.4<br>(69.1-91.6) | 81.3<br>(73.3-87.8) | 100<br>(29.2-100) | 11.1<br>(4.9-20.7) | - |  |
| Specificity, % (95%CI) | 78.5<br>(75.1-81.6) | 86.0<br>(82.9-88.8) | 73.9<br>(70.5-77.2) | 71.8<br>(68.1-75.4) | 93.4<br>(90.7-95.6) |  |
| Anaemia* (baseline), n/N (%) | 20/43 (47) | 41/92 (45) | 3/4 (75) | 15/37 (41) | 120/398 (30) | **0.008** |
| Creatinine** (μmol/L), median (IQR) | 106 (80-150) | 114 (71-177) | 114 (194-194) | 97 (81-132) | 97 (53-300) | 0.995 |
| Platelet count, ×10<sup>3</sup>/μL, median (IQR) | 101<br>(55-138)<br>[12-425] | 97<br>(67-148)<br>[11-565] | 75<br>(56-178)<br>[50-266] | 227<br>(148-309)<br>[11-520] | 237<br>(162-318)<br>[15-610] | **<0.001** |
| Thrombocytopenia, n/N (%)<br>(Platelet count <150×10<sup>3</sup>/μL) | 36/42 (86) | 69/92 (75) | 3/4 (75) | 10/37 (27) | 87/399 (22) | **<0.001** |
| Severe malaria***, n (%)<br>[95%CI] | 0<br>(0) | 1<br>(0.6)[0-3.3] | 0<br>(0) | 0<br>(0) | - | 0.244 |

- Control = participant malaria-negative on PCR with acute febrile illness † Sensitivity/specificity calculated for *Plasmodium* species mono-infection vs non-*Plasmodium* species detection on microscopy against reference PCR ‡ Sensitivity/specificity calculated for PAN-pLDH detection for *P. knowlesi* vs non-*P. knowlesi* on reference PCR \*Anaemia based on WHO 2011 haemoglobin measurement criteria \*\*Creatinine measurement obtained in 36% of malaria cases and 49% of controls \*\*\* Severe Malaria defined using WHO 2014 Research and Epidemiological criteria

There were a single patient meeting 2014 research and epidemiological criteria for WHO-defined severe disease (0.4% of malaria cases with an identified *Plasmodium* species; 95%CI 0.01-2.3%): an adult male with vivax malaria and a haemoglobin of 6.3 g/dL and parasite count of 3,120/µL.

On routine microscopy, the overall sensitivity for *Plasmodium* species detection against reference PCR was 70.1% (95%CI 64.8-75.1%), with a specificity of 95.7% (95%CI 93.8-97.1%) against malaria-negative controls. *P. falciparum* single infections were correctly identified in 42 (59.2%) of patients, with a further 21 cases (29.6%) misidentified as mixed infections. Similarly, single *P. vivax* infections were correctly identified in 62% of participants, however misidentified as mixed *P. vivax* infections in a further 27%. In contrast, there were only small numbers of false-positive microscopy results for *P. falciparum* and *P. vivax* single infections, leading to high diagnostic specificities of 98.4% (95%CI 97.3-99.1%) and 92.4% (95%CI 90.3-94.1%) for each species, respectively. The geometric mean parasite count for *P. falciparum* infections was low at 2,844/μL (95%CI 2,055-3,936/μL), similar to *P. vivax* at 2,273/μL (95%CI 2,487-3,187/μL).

The performance of the combined PAN-pLDH/Pf-HRP2 RDT was poor compared to reference PCR, with a sensitivity and specificity of 82.4% (95%CI 69.1-91.6%) and 78.5% (75.1-81.6%) for *P. falciparum,* and of 81.3% (95%CI 73.3-87.8%) and 86.0% (95%CI 82.9-88.8%) for *P. vivax*. Sensitivity of the RDT compared to microscopy against PCR as the reference was higher overall for detection of *P. falciparum* (82% vs 52%; *p*=0.009) and *P. vivax* (81% vs 62%; *p*<0.001) single infections.

Students were the most frequently reported primary occupation among malaria cases, accounting for ∼50% overall (**Supplemental Table 1**), reflecting the predominantly adolescent cohort. On univariable analysis, a history of sleeping outdoors was associated with higher malaria risk, while insecticide-treated bed net use and personal insect repellent were protective. Housing type also played a key role: concrete walls were protective, while open eaves increased risk. Proximity to mangroves or cleared forest areas was also associated with higher odds of infection. Factors that remained independently associated with malaria in the multivariable analysis are summarised in **Figure 3**.

**Figure 3:**
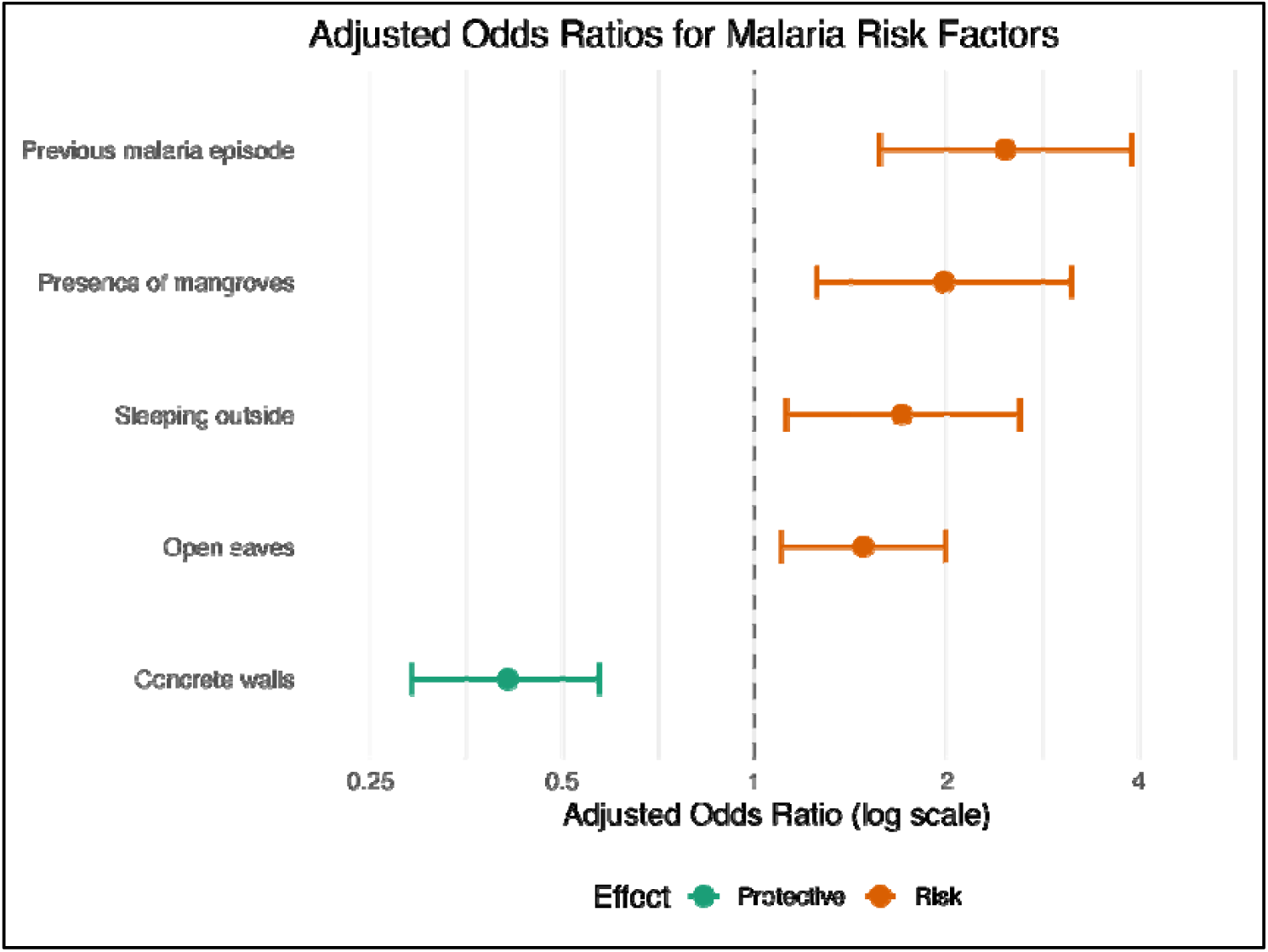
Forest plot of adjusted odds ratios for epidemiological associations with malaria acquisition risk.

### Mursala Island cross-sectional survey

In a small cross-sectional survey conducted on Mursala Island in September 2019, 64 individuals were enrolled (**Table 2**). The median age was 27 years (IQR 15.5–40; range 1– 80), and 58% were female. Most participants (92%) were symptomatic, including 35% reporting a history of fever. PCR detected *Plasmodium* spp. infections in 9 individuals (14.1%, 95% CI: 6.6–25%), comprising 6 *P. knowlesi*, 1 *P. vivax*, 1 mixed *P. knowlesi/P. vivax*, and 1 *Plasmodium* genus-positive case with undetermined species. No infections were detected in patients under 18 years of age. None of the *Plasmodium* species-infected individuals reported a prior malaria diagnosis.

**Table 2.** Clinical and laboratory features of *Plasmodium* species infections in Mursala Island.

| Patient characteristic | North Sumatra – Mursala Island |  |  |  | *P*-value<br>(*Pk* vs *control*) |
| --- | --- | --- | --- | --- | --- |
|  | *P. knowlesi* | *P. vivax* | *P. knowlesi*<br>/ *P. vivax* | *Control* |  |
| Number enrolled, n (%) | 6 (10) | 1 (2) | 1 (2) | 55 (86) | - |
| Age, years |  |  |  |  |  |
| Median | 46 | 34 | 26 | 25 | **0.025** |
| IQR | 30-60 | - | - | 14-40 |  |
| Range | 23-72 | - | - | 1-80 |  |
| Children (age <15 yrs), n (%) | 0 (0) | 0 (0) | 0 (0) | 14 (25) | 0.321 |
| Male gender, n (%) | 3 (50) | 0 (0) | 0 (0) | 24 (44) | 0.999 |
| Previous malaria (self-reported), n (%) | 0 | 0 | 0 | 16 (29) | 0.326 |
| Symptoms on enrolment, n (%) |  |  |  |  |  |
| Any symptom | 6 (100) | 1 (100) | 1 (100) | 50 (91) | 0.999 |
| Fever history (within 48 h) | 2 (30) | 1 (100) | 0 | 31 (9) |  |
| Headache | 6 (100) | 1 (100) | 0 | 42 (12) |  |
| Nausea | 3 (50) | 0 | 1 (100) | 21 (6) |  |
| Cough | 6 (100) | 0 | 0 | 35 (10) |  |
| Shortness of breath | 4 (75) | 1 (100) | 1 (100) | 17 (5) |  |
| Vomiting | 0 (0) | 0 | 0 | 15 (4) |  |
| Abdominal pain | 5 (83) | 1 (100) | 0 | 31 (9) |  |
| Fever (temperature >37.5°C) | 0 | 0 | 0 | 0 | - |
| Febrile days (prior to presentation), n (IQR)[range] | 4.5<br>(2-7)[2-7] | 14 | - | 7<br>(2-7)[1-30] | 0.670 |
| Microscopy result - positive | 0 (0) | 0 (0) | 0 (0) | 0 (0) | - |
| RDT result (PAN-pLDH/Pf-HRP2) | 0 (0) | 0 (0) | 0 (0) | 0 (0) | - |
| Anaemia* (baseline), n/N (%) | 3/3 (100)* | 0/1 (0) | 1/1 (100) | 15/41 (37) | 0.062 |

\*WHO age- and sex-based criteria<sup>38</sup> - includes 2 adult patients with *P. knowlesi* and *P. knowlesi/P. vivax* infections with haemoglobin results of 5.1 g/dL and 4.4 g/dL on Hemocue testing respectively

Overall, 7 patients (10.9%, 95% CI: 4.7–21.9%) were infected with *P. knowlesi,* with a higher median age (36 years [IQR 26–60]) compared to malaria-negative or non-*P. knowlesi* cases (17 years; *p*L=L0.037). Females accounted for 57% of *P. knowlesi* cases. Common symptoms among *P. knowlesi* cases included headache, cough, and abdominal pain, although only 29% reported fever in the preceding 48 hours. The median reported duration of fever was 4.5 days (IQR 2–7) for *P. knowlesi* and 14 days for *P. vivax*. Anaemia, defined by WHO age- and sex-specific thresholds^38^, was present in all *P. knowlesi*-infected individuals tested. Two adults met WHO criteria for severe malaria^30^, with haemoglobin levels of 4.4 g/dL and 5.1 g/dL, respectively. Compared to non-malaria controls, *P. knowlesi* cases had significantly lower median haemoglobin levels (7.1 vs. 13 g/dL; *p*L=L0.004). No other severe malaria features were observed. However, further assessment of respiratory distress or organ dysfunction (e.g., acute kidney injury, hyperbilirubinaemia, hypoglycaemia, metabolic acidosis) was limited by the lack of appropriate clinical and laboratory facilities, respectively. None of the *P. knowlesi* or *P. vivax* infections were detected by microscopy or the combined PAN-pLDH/Pf-HRP2 rapid diagnostic test.

In Mursala, although the small survey size limits meaningful comparisons, five of the seven individuals infected with *P. knowlesi* had occupations related to agriculture (four farmers, one plantation worker; **Supplemental Table 2**). Recent forest exposure (>4 hours) and proximity to rubber plantations were more common among *P. knowlesi* cases (86% each) than controls (46% and 43%, respectively), *p*=0.076 for both comparisons. However, forest activities related to wood collection were associated with higher risk of *P. knowlesi* acquisition (OR 16, 95%CI 1.8-144, p=0.013). Bed net use the night before enrolment was common in both groups (86% of *P. knowlesi* cases vs. 64% of controls). Only one *P. knowlesi*-infected case reported sleeping outside in the previous two weeks. Regular awareness of monkey presence was high among cases (86%), though not significantly different from controls. No household structural or environmental factors, including wall type, elevation, or open eaves, were significantly associated with infection.

## Discussion

This study aimed to further characterise the distribution of zoonotic *Plasmodium knowlesi* infections in North Sumatra, Indonesia using accurate molecular tools. Key findings include the presence of previously unreported *P. knowlesi* submicroscopic symptomatic infections on the remote rural island of Mursala. Additionally, results highlight the significant ongoing burden of non-zoonotic malaria (34%) detected by PCR among acute febrile illness presentations in densely populated mainland coastal areas. North Sumatra public health reporting estimated that *P. falciparum* caused 15% of clinical malaria and *P. vivax* 85% in 2019 (Provincial Health Office 2019, unpublished). In our study, the predominance of *P. vivax* infections underscores the ongoing contribution of this species to the malaria burden despite national elimination efforts, with a doubling of malaria cases reported in North Sumatra since the COVID pandemic in 2020^25^. While Mursala Island had a lower overall malaria prevalence, submicroscopic infections were predominantly caused by zoonotic *P. knowlesi* transmission. No other zoonotic *Plasmodium* species such as *P. cynomolgi* were identified.

Although *P. knowlesi* infections have been previously reported in North Sumatra^9,18^, none were detected during this study among participants from densely populated coastal mainland sites located away from higher-risk agricultural or forest areas. Modelled transmission suitability for *P. knowlesi* in the coastal districts of Batubara and Central Tapanuli is relatively low, with median values of 0.12 (IQR 0.05-0.38) and 0.35 (IQR 0.13-0.55), respectively, compared to 0.61 (IQR 0.27-0.82) in the more interior region of South Tapanuli (unpublished data from Tobin et al^3^). This finding likely reflects distinct epidemiological patterns, similar to those in Malaysian Borneo, where heterogeneous *P. knowlesi* transmission is concentrated in rural areas undergoing land use change^3,39,40^. Unlike Malaysia, however, there is limited understanding about the distribution and behaviour of suspected *Anopheles leucosphyrus* group mosquito vectors across Sumatra^41,42^, or the variation in *P. knowlesi* infection prevalence in local macaque populations^19,42,43^, both of which are key determinants of spatial zoonotic transmission patterns^3,5,44^. However, the demographic profile of *P. knowlesi*-infected individuals - exclusively adults - suggests higher exposure among those engaged in agricultural activities where macaque hosts are present. Both males and females had *P. knowlesi* infections detected, with similar self-reported exposure patterns aligning with findings from other areas of previously reported zoonotic malaria transmission in Malaysia^45^ and elsewhere in western Indonesia^14^. In contrast, *P. falciparum* and *P. vivax* infections in this study predominantly affected younger adolescents, consistent with well-established demographic patterns in other endemic regions approaching elimination of these non-zoonotic species^46^.

The majority of zoonotic and non-zoonotic malaria cases presented with non-specific clinical symptoms, highlighting the ongoing disease burden of malaria and critical need for vigilant point-of-care diagnostic testing. Laboratory findings of moderate anaemia and thrombocytopaenia were more common in malaria-positive patients, which may aid clinical suspicion for diagnostic screening. The small number of patients with *P. knowlesi* infections from Mursala all exhibited anaemia despite extremely low-level submicroscopic parasitaemia, including two cases that met WHO criteria for severe malarial anaemia (haemoglobin <7g/dL)^30^. While chronic causes of anaemia cannot be excluded—particularly given the high baseline prevalence in malaria-negative individuals—*P. knowlesi*-associated anaemia may also result from splenic retention of less-deformable uninfected erythrocytes, and/or bystander destruction of uninfected erythrocytes or dyserythropoiesis^47,48^ , potentially exacerbating underlying vulnerability. Evidence from Malaysia similarly reports anaemia in uncomplicated *P. knowlesi* infections, although generally less prevalent and severe than in *P. falciparum* and *P. vivax* malaria^49,50^. The low parasitaemia levels in Mursala cases could reflect partial immunity from repeated *P. knowlesi* exposure, cross-protection from prior *P. vivax* infections, or reduced erythrocyte invasion due to parasite or host genetic factors, including haemoglobinopathies^51^. Parasite counts of >15,000/μL have been linked to a >16-fold risk of severe *P. knowlesi* malaria in Malaysian Borneo, often with clinical complications such as jaundice, respiratory distress, hypotension, and acute kidney injury^49,50^. These manifestations were not observed either due to the low parasite counts or the limited laboratory testing available in this study.

The threat of emerging *P. knowlesi* transmission highlights the ongoing importance of access to robust diagnostic capabilities and antimalarial treatment in provinces approaching elimination of non-zoonotic *Plasmodium* species. Indonesia’s national malaria control programme currently relies on passive surveillance using microscopy or PAN-pLDH/Pf-HRP2-based RDTs^52^. In our study, microscopy demonstrated only moderate sensitivity for detecting *P. falciparum* (59.2%) and *P. vivax* (62.0%) mono-infections, with notable difficulty distinguishing mixed infections. This raises concerns around misclassification of *P. vivax* which may result in a lack of primaquine administration for radical liver cure. The PAN-pLDH/Pf-HRP2 RDT showed strong performance in detecting *P. falciparum* and *P. vivax*, reinforcing its utility in field settings. However, our molecular diagnostics identified a high proportion of submicroscopic (30%) and *Plasmodium* genus (undetermined species) infections (25%) in mainland sites, likely due to the enhanced sensitivity of the reverse transcriptase real-time PCR workflow with a lower limit of detection in the *Plasmodium* genus compared to species-specific assays^33^. However, we cannot exclude the possibility of false-positive results arising from sample inhibitors, degraded RNA, or other assay-related factors. In contrast, conventional point-of-care tests failed to detect the low-density *P. knowlesi* infections found in patients on Mursala Island despite their symptomatic presentation. This aligns with previous studies showing poor performance of PAN-pLDH-based RDT for *P. knowlesi*^11,12^, although the sensitivity of more recent RDTs has improved for parasite counts greater than 200/µL^10^. These limitations emphasise the critical need to integrate molecular tools into surveillance systems in areas approaching malaria elimination, especially where *P. knowlesi* is suspected^28^.

*Plasmodium knowlesi* infection has been associated with environmental and occupational risk factors, including forest and agricultural work (particularly among farmers and plantation workers), sleeping outside, and household characteristics such as open eaves or gaps in walls^45^. In our study, while the majority of *P. knowlesi* cases (71%) were engaged in agricultural activities, this occupation alone did not significantly increase infection risk. Recent forest exposure, house proximity to rubber plantations, and wood collection in forested areas were identified commonly for the small number of *P. knowlesi* infections in Mursala Island, although we were underpowered to determine if these were also statistically significant associations in this setting. It is likely these trends indicate a zoonotic transmission cycle involving forest-dwelling vectors and macaque reservoirs, contrasting with the coastal semi-urban environments of our mainland study sites where *P. knowlesi* infections were not detected. Furthermore, while malaria prevention measures such as bed net use and personal insect repellent use may continue to provide benefits for preventing non-zoonotic malaria, their use did not provide significant protection against *P. knowlesi* infection in our study. This finding emphasises the need for tailored prevention strategies targeting high risk groups engaged in forest-related activities to effectively mitigate zoonotic malaria transmission risk.

Our study has several limitations. In the mainland study sites, there was an over-representation of young individuals, with only 14% of participants being adult men aged over 30 years, in addition to only 3% of all participants reporting either agricultural occupations or recent forest exposure. This might have introduced a degree of selection bias by insufficiently capturing high-risk populations, particularly adult forest and agricultural workers, who are most vulnerable for zoonotic malaria^45^ . The study’s design focus on symptomatic cases presenting to health facilities potentially underestimates the true infection burden by missing mild and asymptomatic cases that did not seek medical attention^53^. Additionally, the COVID-19 pandemic interrupted both the study timeline and routine malaria activities, potentially affecting healthcare-seeking behaviour and contributing to increased malaria burden during the study period. On Mursala Island, the cross-sectional nature of the surveys and relatively small number of *P. knowlesi* cases limit our ability to draw definitive conclusions about risk factors and transmission patterns. Furthermore, the lack of laboratory facilities for comprehensive assessments of WHO-severity criteria prevented thorough evaluation of key parameters of severe malaria, particularly in *P. knowlesi* cases.

## Conclusion

Our study highlights the complex malaria epidemiology in North Sumatra and the ongoing challenges to Indonesia’s elimination goals. Despite control efforts, non-zoonotic malaria— particularly *P. vivax*—remains a major cause of febrile illness on the mainland. In contrast, *P. knowlesi* transmission on Mursala Island reflects distinct zoonotic dynamics, linked to forest-related activities such as rubber plantation proximity, wood collection, and recent forest travel. These findings underscore the need to strengthen molecular diagnostic capacity and adopt improved sampling strategies in future surveillance to better understand and mitigate the growing threat of zoonotic malaria in Indonesia.

## Data Availability

All data produced in the present study are available upon reasonable request to the authors.

## Acknowledgements

We thank the study participants, and the health facility and research staff involved in the data collection.

This work was supported by the Australian Centre for International Agricultural Research and the Department of Foreign Affairs and Trade, Australian Government (LS/2018/214), and the National Health and Medical Research Council, Australia (fellowships to NMA [1042072], MJG [1138860]).

## Author contributions

INL, BEB, NMA, and MJG conceived the study design; RP, LS, RADC, IRAN, RDS, MNS, SJ, and AL performed data collection and conducted the experiments; INL, IRAN and MJG performed data analysis and wrote the original draft; BEB, KAP and NMA provided supervision, funding acquisition and resources; All authors read, reviewed and approved the final manuscript.

## Author Bio

Dr Inke Lubis is a paediatrician and clinical researcher at Universitas Sumatera Utara, Indonesia, with extensive experience in malaria epidemiology, antimalarial resistance markers, and One Health approaches to infectious disease control. She has been actively involved in supporting Indonesia’ National Malaria Control Programme through her research and policy engagement.

