## Supplemental Tables for "Submicroscopic burden of zoonotic *P. knowlesi* malaria on Mursala Island and *P. falciparum* and *P. vivax* transmission in mainland North Sumatra, Indonesia"

**Supplemental Table 1 –** Epidemiological factors associated with *P. falciparum* and *P. vivax* infections on mainland sites

| **Patient characteristic** | **North Sumatra – mainland sites** | | | | | |
| --- | --- | --- | --- | --- | --- | --- |
|  | ***P. falciparum*** | ***P. vivax*** | ***P. falciparum /P. vivax*** | ***Plasmodium* genus** | ***Control***  *(negative)* | ***P-value***  *(across groups)* |
| Number enrolled | 71 | 166 | 5 | 79 | 626 |  |
| ***Occupation*** |  |  |  |  |  |  |
| Agriculture | 2 (2.8) | 5 (3.1) | 0 (0) | 2 (2.5) | 25 (4.0) | 0.908 |
| Fisherman | 7 (9.9) | 8 (4.9) | 0 (0) | 0 (0) | 28 (4.5) | 0.071 |
| Household/family carer | 7 (9.9) | 16 (9.8) | 0 (0) | 11 (13.9) | 93 (14.9) | 0.328 |
| No work/unemployed | 7 (9.9) | 13 (7.9) | 0 (0) | 10 (12.7) | 66 (10.6) | 0.713 |
| Other | 10 (14.1) | 25 (15.2) | 3 (60) | 7 (8.9) | 100 (16.0) | **0.032** |
| Self-employed/business | 2 (2.8) | 8 (4.9) | 0 (0) | 5 (6.3) | 50 (8.0) | 0.342 |
| Student | 36 (36) | 89 (54.2) | 2 (40) | 44 (55.7) | 262 (42.0) | **0.016** |
| ***Activities*** |  |  |  |  |  |  |
| Sleep outside house  (last 2 weeks) | 11 (15.5) | 33 (20.1) | 0 (0) | 9 (11.4) | 58 (9.3) | 0.003 |
| Bednet use outside house | 1 (1.4) | 5 (3.1) | 0 (0) | 2 (2.5) | 6 (1.0) | 0.334 |
| Aware of monkeys | 31 (43.7) | 59 (36.0) | 3 (60) | 16 (20.3) | 193 (30.9) | **0.012** |
| Clearing forest/vegetation | 8 (11.3) | 10 (6.1) | 0 (0) | 7 (8.9) | 32 (5.1) | 0.218 |
| Forest exposure (>4 hours) | 4 (5.6) | 5 (3.1) | 1 (20) | 4 (5.1) | 14 (2.2) | 0.058 |
| Cutting timber | 0 (0) | 0 (0) | 0 (0) | 0 (0) | 2 (0.3) | 0.906 |
| Collecting wood | 0 (0) | 2 (1.2) | 0 (0) | 0 (0) | 4 (0.6) | 0.764 |
| Hunting | 0 (0) | 0 (0) | 0 (0) | 0 (0) | 1 (0.2) | 0.972 |
| Charcoal making | 0 (0) | 0 (0) | 0 (0) | 1 (1.3) | 1 (0.2) | 0.313 |
| Other forest activity | 1 (1.4) | 3 (1.8) | 0 (0) | 0 (0) | 3 (0.5) | 0.378 |
| ***Malaria prevention*** |  |  |  |  |  |  |
| Bed net use (any) | 51 (71.8) | 120 (73.2) | 5 (100) | 59 (74.7) | 419 (64.1) | 0.197 |
| Bed net use  (Insecticide treated) | 19 (26.8) | 35 (21.3) | 2 (40) | 16 (20.3) | 179 (28.3) | 0.210 |
| Personal insect repellent  (lotion/spray) | 9 (12.7) | 22 (13.4) | 0 (0) | 6 (7.6) | 114 (18.3) | 0.064 |
| ***Household*** |  |  |  |  |  |  |
| Wall construction |  |  |  |  |  |  |
| Wood | 18 (25.3) | 50 (30.5) | 1 (20) | 45 (57.0) | 185 (29.7) | **<0.001** |
| Concrete | 13 (18.3) | 34 (20.7) | 1 (20) | 14 (17.7) | 253 (40.5) | **<0.001** |
| Tin | 1 (1.4) | 3 (1.8) | 0 (0) | 2 (2.5) | 1 (0.2) | 0.052 |
| Bricks | 30 (42.3) | 67 (40.9) | 3 (60) | 17 (21.5) | 164 (26.3) | **<0.001** |
| Bamboo | 5 (7.0) | 0 (0) | 0 (0) | 0 (0) | 3 (0.5) | **<0.001** |
| Other | 4 (5.6) | 10 (6.1) | 0 (0) | 1 (1.3) | 18 (2.9) | 0.179 |
| Elevated (>1m stilts) | 19 (26.8) | 18 (11.0) | 1 (20) | 19 (24.1) | 121 (19.4) | **0.026** |
| Open eaves/gaps | 35 (49.3) | 85 (51.8) | 2 (40) | 34 (43.0) | 203 (32.5) | **<0.001** |
| Insecticide treated walls (IRS) | 4 (5.6) | 8 (4.9) | 0 (0) | 3 (3.8) | 32 (5.1) | 0.964 |
| ***Household environment (±100m)*** |  |  |  |  |  |  |
| Oil palm | 28 (39.4) | 57 (34.8) | 3 (60) | 19 (24.1) | 215 (34.5) | 0.199 |
| Rubber | 1 (1.4) | 7 (4.3) | 0 (0) | 0 (0) | 18 (2.9) | 0.367 |
| Rice paddy | 3 (4.2) | 18 (11.0) | 3 (60) | 4 (5.1) | 55 (8.8) | **<0.001** |
| Mangrove | 14 (19.7) | 25 (15.2) | 2 (40) | 6 (7.6) | 48 (7.7) | **<0.001** |
| Cleared forest area | 9 (12.7) | 9 (5.5) | 2 (40) | 3 (3.8) | 20 (3.2) | **<0.001** |
| Intact forest area | 13 (18.3) | 16 (9.8) | 2 (40) | 5 (6.3) | 13 (12.8) | **0.046** |

**Supplemental Table 2 –** Epidemiological factors associated with *P. knowlesi* infections on Mursala Island

| **Patient characteristics** | ***P. knowlesi*** | ***Control***  *(malaria negative)* | ***Odds ratio***  *(95% CI)* | ***P-value*** |
| --- | --- | --- | --- | --- |
| Number enrolled | 7 | 55 |  |  |
| ***Occupation*** |  |  |  |  |
| Agriculture | 5 | 22 | 4.35 (0.47-40.4) | 0.196 |
| Fisherman | 1 | 10 | 1.82 (0.01-31.9) | 0.683 |
| Unemployed | 1 | 19 | Reference | **-** |
| ***Activities*** |  |  |  |  |
| Sleep outside house  (last 2 weeks) | 1 (14.3) | 14 (25.5) | 0.49 (0.05-4.41) | 0.523 |
| Bednet use outside house | 1 (14.3) | 0 (0) | 3.42 (0.38-30.5) | 0.270 |
| Aware of monkeys | 6 (85.7) | 39 (70.9) | 2.46 (0.27-22.1) | 0.421 |
| Clearing forest/vegetation | 0 (0) | 11 (20) | - | 0.334 |
| Forest exposure (>4 hours) | 6 (85.7) | 25 (45.5) | 7.13 (0.81-63.9) | 0.076 |
| ***Malaria prevention*** |  |  |  |  |
| Bed net use (any) | 6 (85.7) | 35 (63.6) | 3.43 (0.38-30.5) | 0.270 |
| Personal insect repellent use  (lotion/spray) | 0 (0) | 2 (3.6) | - | 0.608 |
| ***Household*** |  |  |  |  |
| Wooden walls | 7 (100) | 51 (92.7) | - | 0.999 |
| Elevated (>1m stilts) | 4 (57.1) | 20 (36.4) | 2.33 (0.47-11.49) | 0.298 |
| Open eaves/gaps | 4 (57.1) | 40 (72.7) | 0.50 (0.10-2.50) | 0.399 |
| Insecticide treated walls (IRS) | 0 (0) | 4 (7.3) | - | 0.461- |
| ***Household environment (±100m)*** |  |  |  |  |
| Oil palm | 0 (0) | 0 (0) | - | - |
| Rubber | 6 (85.7) | 25 (42.5) | 7.20 (0.81-63.9) | 0.076 |
| Rice paddy | 0 (0) | 1 (1.8) | - | 0.719 |
| Mangrove | 2 (28.6) | 13 (23.6) | 1.29 (0.22-7.46) | 0.774 |
| Cleared forest area | 1 (14.3) | 12 (21.8) | 0.60 (0.07-5.45) | 0.648 |
| Intact forest area | 7 (100) | 36 (65.5) | - | 0.062 |
